# Ocular findings, surgery details and outcomes in proliferative diabetic retinopathy patients with chronic kidney disease

**DOI:** 10.1101/2022.08.04.22278408

**Authors:** Jipeng Li, Aman Chandra, Lin Liu, Lin Zhang, Jun Xu, Meng Zhao

## Abstract

**Purpose:** We investigated the influence of impaired renal function on fundus characteristics, pars-plana vitrectomy (PPV) details, and outcomes in patients with proliferative diabetic retinopathy (PDR).

**Design:** a retrospective cohort study

**Methods:** We investigated a consecutive series of PDR patients who underwent PPV. The diabetic complications, previous photocoagulation, intravitreal injections before PPV, ocular findings during PPV, surgical details, short-term visual outcome and post-PPV complications were recorded and compared between patients with and without impaired renal function.

**Results:** 149 patients had normal renal function (67.7%) and 71 (32.3%) patients had impaired renal function, 85.4% of patients were identified with chronic kidney disease (CKD) during the preoperative assessment. Impaired renal function was related to hypertension (3.40[1.58-7.29], p=0.002), incomplete panretinal photocoagulation (PRP) (3.18[1.50-6.72], p=0.002), severe fibrovascular membrane (8.19[3.43-19.54], p<0.001), and extensive retinal vascular closure (3.40[1.54-7.52], p=0.002). There was a more frequent occurrence of severe intraoperative bleeding (56.3%, 32.2%, p=0.001), a higher percentage of intraocular subretinal fluid drainage (45.1%, 22.1%, p=0.008) in patients with impaired renal function. The percentage of patients whose visual acuity (VA) increased was similar between two groups (46.4%, 54.3%, p=0.34).

**Conclusions:** In PDR patients, screening for CKD was required before PPV. PDR patients with impaired renal function tended to have more severe ischemic retinal conditions. Comparable PPV outcomes could be obtained in patients with and without impaired renal damage.

## Background

Proliferative diabetic retinopathy (PDR) and chronic kidney disease (CKD) are microvascular complications of diabetes mellitus (DM), with prevalence rates of 30-40%^1^ and 18.45%^2^, respectively. The two complications often coexist in patients with long-standing DM, and the prevalence of patients with an impaired estimated glomerular filtration rate (eGFR) in patients with diabetic retinopathy (DR) is 21.3-40.9%^3,4^; conversely, the prevalence of DR in patients with an impaired eGFR is 26.7-38.5%^4,5^.

Fundus photography^6-8^ is the standard screening tool for DR severity evaluation and can be of little value when the fundus of a patient is obscured by vitreous hemorrhage (VH) or tractional retinal detachment (tRD). The two conditions are commonly anticipated to be severe complications of PDR. PDR patients with VH or tRD may require pars plana vitrectomy (PPV). The PPV provides the opportunity to investigate obscured fundus characteristics. The previous work on PDR patients who underwent PPV implies that patients with impaired renal function can achieve a similar visual outcome with tolerable postoperative complications ^9,10^, but renal failure is the leading cause of mortality in the follow-up^11^. However, there was little information on the fundus characteristics and operation details in previous work on PDR patients with CKD who underwent PPV.

The prevalence of DM in China has increased 17-fold from 0.67% to 11.6% of the population in the past 30 years^12^. The prevalence of CKD in China is increasing^13^. Dialysis has provided PDR patients with end-stage renal failure opportunities for PPV^10,11^. Therefore, ophthalmologists must face the challenge of performing PPV on PDR patients with CKD. Herein, we investigated a group of PDR patients who underwent PPV, and we compared the fundus characteristics, operative details, and PPV outcomes between patients with impaired renal function and patients with normal renal function.

## Method

This study was a retrospective cohort of consecutive hospitalized patients with PDR who underwent PPV at our hospital during 2016.1.1-2017.12.31. The eye with more severe ocular manifestations was selected if there was bilateral involvement. The authors had access to information that could identify individual participants during or after data collection.

### Patient enrollment and grouping

#### 1) Inclusion and exclusion criteria

Inclusion criteria: 1) PDR patients who have been hospitalized for PPV because of a nonclearing VH (persistent or recurrent) or tRD; 2) the high-risk PDR ^14^defined by Early Treatment of Diabetic Retinopathy Study (EDTRS) guidelines were confirmed in operation; 3) available history records and laboratory tests; 4) detailed operation records; 5) detailed follow-up records for at least three months.

The exclusion criteria were as follows: patients with one or more of the following conditions were excluded from the study: 1) vitreous hemorrhage not related to PDR during vitrectomy; 2) eyes with PDR that had PPV for the epiretinal membrane or macular hole; 3) eyes with a history of PPV; 4) a lack of history of DM, or lack of diagnosis information for hypertension (HBP), DM-related cardiocerebral vascular disease or CKD; 5) a lack of operation data for further statistical analysis; 6) a lack of 3-month follow-up; or 7) a history of other renal diseases that caused abnormal renal function.

#### 2) Grouping based on stage of CKD

CKD was defined as diabetes with albuminuria, impaired eGFR, or both according to the National Kidney Foundation KDOQI (Kidney Disease Outcomes Quality Initiative) classification^19^. We classified CKD into the following ranges by eGFR: stage 1 (≥ 90 ml/min/1.73 m^2^); stage 2 (60–89 ml/min/1.73 m^2^); stage 3 (30–59 ml/min/1.73 m^2^); stage 4 (15–29 ml/min/1.73 m^2^); and stage 5 (<15 ml/min/1.73 m^2^). Impaired renal function was defined as eGFR<60 ml/min per 1.73 m^2^ (stage 3 or worse than stage 3) ^20^.

All patients were divided into two groups: patients with normal renal function (CKD stage 1-2) and patients with impaired renal function (CKD stage 3-5).

### Baseline investigations

#### 1) Systemic complication investigation

Baseline data were collected from past history and presurgical anesthesia assessment records. The following information was included: age, sex, DM duration, DM medication, history of diabetes-related complications, including stroke, coronary heart disease (CAD), congenital heart failure (HF), diabetic foot, CKD, HTN. All the patients underwent laboratory tests on blood cell counts, urine analysis, blood coagulation function tests, serum creatinine, and blood urine nitrogen. All patients underwent a preoperative routine 12-lead electrocardiogram.

#### 2) The ocular findings

The history of previous panretinal photocoagulation (PRP), cataract phacoemulsification extraction, and intravitreal injection (IV) of anti-vascular endothelial growth factor (VEGF) agents was recorded. The course of unresolving VH from the onset of symptoms to the presentation, recurrent or persistent, was recorded.

All patients underwent comprehensive ophthalmological examinations, including corrected visual acuity (VA), which was tested using a decimal VA chart, slit-lamp biomicroscopy, intraocular pressure (IOP) measurement, dilated fundus examination with indirect ophthalmoscopy, color fundus photograph, optical biometry, coherent optic tomography (OCT) and B scan. Corrected bilateral VA, iris neovascular or neovascular glaucoma and dense vitreous hemorrhage in the operated eye were recorded.

### Operative ocular findings and management

All patients underwent 23/25 gauge 3-port PPV (Constellation Vision System, Alcon, Fort Worth, Texas, USA) (Stellaris PC, Bausch+LOMB, USA) with 5000 cuts/min vitreous cutting rates. Patients with active fibrovascular epiretinal proliferative membranes (FVPs) or severe VH obscured fundus were treated with intravitreal injection of ranibizumab (IVR) within seven days before PPV. Pre-PPV IVR was recorded.

After core vitrectomy to remove opacified vitreous, the extent of the FVP, the presence of macular-involving TRD, photocoagulation scars and the retinal vessel status were recorded.

IV triamcinolone acetonide (TA)-assisted PPV was performed. Anterior-posterior vitreoretinal traction was released as much as possible. Induction of PVD, FVP dissection, segmentation, delamination, endodiathermy and drainage of subretinal fluid, retinectomy and intraocular tamponade were performed as required according to each subject’s particular need. Peripheral vitrectomy with scleral indentation and endolaser photocoagulation were performed in each case. IOP elevation, perfluorocarbon liquids and endodiathermy were used to handle the intraoperative bleeding. The severity of intraoperative bleeding was recorded as follows:

grade 0: none;

grade 1: minor bleeding stopping spontaneously or with transient bottle pressure elevation;

grade 2: moderate to severe bleeding requiring endodiathermy or with the formation of broadsheets of clots^15^.

Incomplete scatter photocoagulation was defined as a lack of preexisting photocoagulation in all four quadrants, fewer than 1000 laser spots or the required additional more than 500 laser spots during PPV^16^.

The operation time was calculated from making the first PPV trocar incision to removing the eyelid speculum after the PPV. The combined procedures were recorded, including cataract extraction, silicone oil tamponade, photocoagulation, and the use of endodiathermy. The laser points were recorded. The extension of FVPs was recorded as follows:

grade 0: absence of any adhesion;

grade 1: multiple point adhesions with or without one broad adhesion (broad adhesion was defined as focal adhesion at three sites or more);

grade 2: 1-3 broad adhesions posterior to the equator;

grade 3: 3+ broad adhesions posterior to the equator or two or fewer in quadrants adhesions anterior to the equator;

Grade 4: broad adhesions anterior to the equator in multiple sites^17,18^.

FVPs were classified as predominantly neovascular, mixed neovascular and fibrotic, and predominantly fibrotic^15^. The marked reduction in the caliber of retinal vessels and widespread retinal vessel closure were recorded.

### Follow-up

All patients were followed for at least three months monthly. All patients underwent corrected VA, slit-lamp biomicroscopy, IOP measurement, and dilated fundus examination with indirect ophthalmoscopy. The VA result, occurrence of reoperated postoperative vitreous hemorrhage (POVH), retinal detachment, and development of neovascular glaucoma (NVG) were recorded. Optic Coherent tomography (OCT) evaluation of macular attachment was carried out on patients with silicone oil tamponade just before undergoing silicone oil removal. The status of the macula was recorded as attached or detached.

### Statistical analysis

Statistical analysis was performed using R version 3.20 (http://www.R-project.org). Patient characteristics were retrieved from their medical charts and recorded in Epidata Entry Client version 2.0.3.15 (http://epidata.dk). The corrected VA results were converted to logMAR values for statistical analysis. The mean and standard deviation (SD) were calculated for continuous variables with a normal distribution.

The median with quartiles was calculated for continuous variables with a nonnormal distribution. The t test or Mann–Whitney U test was carried out for continuous variables. The chi-square test or Fisher’s exact test was carried out for discrete data. To investigate the relationship between impaired renal function and PDR characteristics, the patients were divided into two groups: those with impaired renal function and those with normal renal function. The baseline systemic condition, presurgical findings, and ocular findings during PPV were compared between the two groups. Variables with a p value less than 0.3 were further enrolled in a binary backward stepwise logistic regression model. One variable was included or excluded from the model each time by comparing the Akaike information criterion (AIC) value; the model that had the lowest AIC was chosen.

To investigate the influence of CKD on the PPV and PPV outcomes, the variables of PPV management, VA, retinal attachment rate, and the occurrence of POVH and NVG were compared between the two groups.

## Results

There were 220 patients enrolled in our study with an average age of 51.8±11.8 years. Among them, 126 (57.3%) were males, and 94 (42.7%) were females. There was 5 cases excluded due to failing to follow-up and all of them had stage 1 CKD.

There were 149 patients with stage 1-2 CKD (67.7%) and 71 (32.3%) patients with impaired renal function and stage 3-5 CKD (Figure 1).

**Figure 1.**
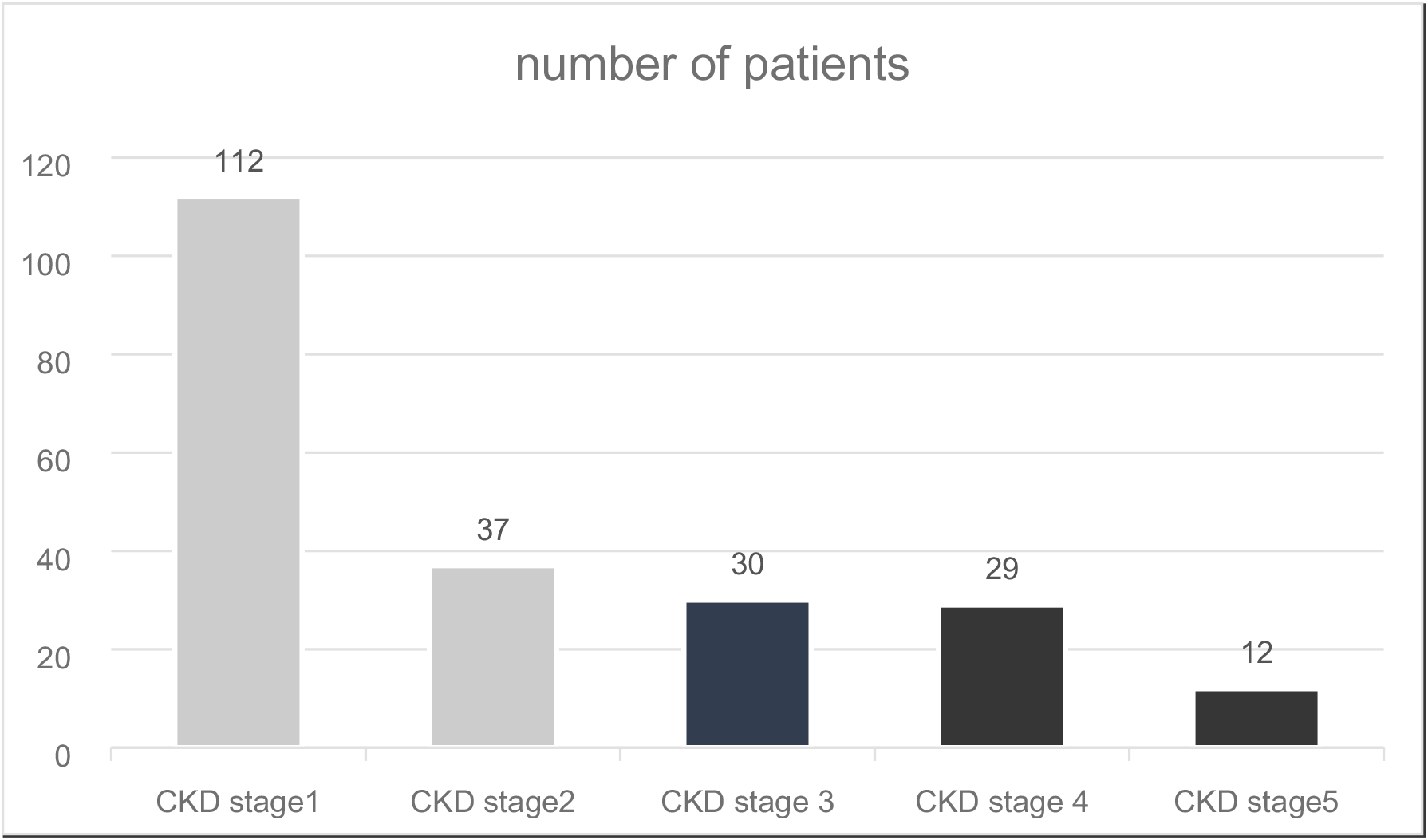
The prevalence of CKD in the investigated PDR patients.

There were 20 (9.1%) patients who had gone through urine examination or renal function test before the onset of ocular symptoms, 12 (5.5%) patients under dialysis, and 188 patients (85.4%) who did not have CKD screening before the preoperative assessment. In addition to the 12 patients undergoing dialysis, four received medical treatment to protect renal function.

### Impaired renal function-related systemic and ocular findings

1. Univariable relationship between impaired renal function and systemic and ocular findings The association of the medical history and preoperative and intraoperative factors with impaired renal function was listed in Table 1. The univariable analysis showed that compared to patients with normal renal function, patients with impaired renal function had a higher percentage of males (70.4%, 51.0%, p<0.001), a higher percentage of coexistent HTN (69.0%, 42.3%, p<0.001), more patients with stroke (9,7, p=0.04), a higher percentage of patients who had incomplete PRP (64.8%, 42.3%, p<0.001), a higher percentage of patients who presented with predominantly fibrotic FVP (43.7%, 25.5%, p=0.01), a higher percentage of patients who presented with board FVP (>= stage 3) (77.5%, 28.9%, p<0.001), a higher percentage of patients with macula-involved tRD (46.5%, 24.1%, p<0.001), and a higher percentage of patients with extensive retinal vessel closure (63.3%, 13.4%, p<0.001).

**Table 1.** The baseline and intraoperative characteristics of PDR patients by the presence of impaired renal function
2. Multiple variables logistics regression Logistic regression showed that factors related to impaired renal function were the presence of HTN (3.40[1.58-7.29], p=0.002), preoperative incomplete PRP (3.18[1.50-6.72], p=0.002), presence of grade 3 FVP (8.19[3.43-19.54], p<0.001), and presence of extensive retinal vascular closure (3.40[1.54-7.52], p=0.002) (AIC=203.4, AUC=0.854).

|  | Patients with<br>impaired renal<br>function (71) | Patients with<br>normal renal<br>function (149) | P |
| --- | --- | --- | --- |
| age (mean±SD, y) | 51.0±13.3 | 52.1±11.1 | 0.55 |
| gender (male,<br>n, %) | 50, 70.4% | 76, 51.0% | <0.001 |
| duration of DM<br>(median, IQR, m) | 120[66,180] | 120[60,208] | 0.27 |
| combined systemic abnormalities |  |  |  |
| combined HTN<br>(n, %) | 49,69.0% | 63,42.3% | <0.001 |
| combined<br>coronary heart<br>disease (n, %) | 20, 28.2% | 47, 31.5% | 0.32 |
| combined stroke<br>(n, %) | 9, 12.7% | 7, 4.7% | 0.04 |
| presurgery ocular findings and ocular medical history |  |  |  |
| VA<0.01(n, %) | 40 | 87 | 0.89 |
| recurrent or<br>persistent VH<br>(n, %) | 53,74.6% | 50,33.6% | <0.001 |
| IVR after VH<br>(n, %) | 3, 6.5% | 5,7.5% | 0.72 |
| previous history of PRP |  |  |  |
| incomplete PRP<br>(n, %) | 46, 64.8% | 63, 42.3% | <0.001 |
| without PRP<br>(n, %) | 22, 31.0% | 82, 55.0% | 0.001 |
| complete PRP (n) | 3 | 3 | 0.68 |
| surgical characteristics (n, %) |  |  |  |
| presence of<br>predominantly<br>fibrotic FVP | 31, 43.7% | 38, 25.5% | 0.01 |
| grade of extension of FVP (n, %) |  |  |  |
| <=grade2 | 16, 22.5% | 106, 71.1% | <0.001 |
| >=grade3 | 55, 77.5% | 43, 28.9% |  |
| macula-involved<br>TRD (n, %) | 33, 46.5% | 36, 24.1% | <0.001 |
| extensive retinal<br>vessel closure<br>(n, %) | 45, 63.3% | 30,13.4% | <0.001 |
| combined retinal<br>tear (n, %) | 26, 32.9% | 56, 39.7% | 0.99 |

### The influence of impaired renal function on PPV

There was no significant difference in the operation time, percentage of patients who received a preoperative IV anti-VEGF agent injection, rate of patients who underwent combined phacoemulsification cataract extraction, number of laser points during the surgery, or percentage of silicone oil tamponade between the two groups (Table 2). There was a more frequent occurrence of grade 2 intraoperative bleeding (56.3%, 32.2%, p=0.001), a higher percentage of intraocular subretinal fluid drainage (45.1%, 22.1%, p=0.008), and a higher percentage of perfluorocarbon liquids (46.5%, 30.9%, p=0.035) in patients with impaired renal function than in patients with normal renal function.

**Table 2.** The surgical characteristics of PDR patients by the presence of impaired renal function

|  | Patients with impaired renal function (71) | Patients with normal renal function (149) | P |
| --- | --- | --- | --- |
| IV-anti VEGF before surgery (n, %) | 58, 81.7% | 132, 88.6% | 0.23 |
| operative time (median, IQR, min) | 95.0[60,120] | 100[60,120], | 0.24 |
| combined cataract extraction (n) | 6 | 7 | 0.46 |
| combined subretinal fluid drainage (n, %) | 32, 45.1% | 33, 22.1% | 0.008 |
| severe intraoperative bleeding (grade 2, n, %) | 40, 56.3% | 48, 32.2% | 0.001 |
| use of perfluorocarbon liquids (n, %) | 33, 46.5% | 46, 30.9% | 0.035 |
| laser points (median, IQR, n) | 980[696,1306] | 1092[669,1595] | 0.12 |
| silicone oil tamponade (n, %) | 24, 33.8% | 50, 33.5% | 0.99 |

### Influence of impaired renal function on PPV outcomes

During a median follow-up of 29 weeks, ranging from 24 to 40 weeks, 46.4% of patients with impaired renal function had VA increased for more than two lines at their 3-month follow-up, similar to the 54.3% of patients with normal renal function (p=0.34); 42.2% of patients with impaired renal function had stable (VA changes were less than two lines) 3-month follow-up VA, similar to the 56.4% patients with normal renal function (p=0.07).

In 74 patients with silicone oil tamponade, macular reattachment was found in all patients at silicone oil removal. Four patients with impaired renal function and three patients with normal renal function had persistent macular edema. In 220 patients, two patients with impaired renal function and one with normal renal function had secondary PPV for recurrent retinal detachment; the retinal attachment rates were 97.2% and 99.3%, respectively.

Four patients with impaired renal function and eight with normal renal function had POVH (p=0.99). POVH occurred after restarting antiplatelet therapy.

No patient in our study developed NVG during the follow-up period.

## Discussion

Previous investigations on the relationship between DR and CKD were carried out in DM patients with transparent optic media. These studies often excluded PDR patients with VH due to failure to obtain a clear fundus photograph. We enrolled PDR patients with VH who underwent PPV and investigated the fundus characteristics of PDR patients after removing the VH by PPV. We found that PDR patients with impaired renal function had specific FVP and retinal vascular characteristics. Correspondingly, specific operative details were different between the two groups. Finally, after proper operative management, we found that PDR patients with impaired renal function could achieve comparable operative outcomes compared with PDR patients with normal renal function.

### Low awareness of CKD in PDR patients who underwent PPV

The prevalence of CKD and impaired renal function in our PDR patients was 49.1% and 32.8%, respectively, far beyond the hospital-based CKD prevalence of 10.7%^19^ and the population-based CKD prevalence of 11.98-29.60%^20,21^ in China. Previous studies showed that the five-year survival rate of PDR patients who underwent PPV was 81-86%^28,29^, and renal failure was the leading cause of mortality in PDR patients who underwent PPV^14,28,29^. The high prevalence of patients with impaired renal function in our study indicated that the patients enrolled in our study were a group of patients with a risk of post-PPV mortality if the renal condition was left untreated.

We further found that among the 220 patients, 85.4% did not know they had CKD, and only 1.8% of patients were under treatment in addition to patients undergoing dialysis. The low awareness rate and treatment rate of CKD in our group of patients were similar to the results of previous population-based investigations in China, with an awareness rate of CKD of 8.7-9.5% and a treatment rate of 4.9%^22-24^. Since most patients with CKD are asymptomatic, our results suggested that ophthalmologists should pay extra attention to the screening and treatment of CKD when dealing with fundus abnormalities in PDR patients who underwent PPV.

### The impaired renal function-related fundus characteristics of PDR

The 4-grade classification system of FVPs based on the severity of vitreoretinal adhesion is related to PPV visual prognosis^17,18^. It also reflects the difficulty in dealing with FVPs, for example, combined retinal tears are often found in patients with grade 3 or more severe FVPs^18^, and retinectomy is often required in patients with grade 4 FVPs^17,18^.

The progression of PDR is related to the diabetic course^25^ and the combination of CKD^26,27^. We divided the patients into two groups: a group with normal renal function (including patients with stage 1-2 CKD) and a group with impaired renal function (including patients with stage 3-5 CKD). There was no significant difference in the duration of the diabetes course between the two groups (120, 120 months). For a similar duration of diabetes, we found that PDR patients with impaired renal function had a higher percentage of patients with severe FVP (grade 3 or more severe) (77.5%, 28.9%), macular-involved tRD (46.5%, 24.1%), extensive retinal vessel closure (63.3%, 13.4%), predominantly fibrotic FVP (43.7%, 25.5%), and incomplete PRP (64.8%,42.3%) than PDR patients with normal renal function. Previous research in patients with clear optic media shows that large nonperfusion areas in DM patients are related to CKD progression ^8^, while the combination of CKD is related to DR progression^26,27^. We extended the relationship of CKD and DR to patients who underwent PPV with unclear optic media, and our results showed that impaired renal function was related to more severe ischemic retinal changes, including retinal vessel closure and board FVP. It suggests that more aggressive treatment for CKD in patients with DR should be applied, and more attention should be given to DR progression in CKD patients.

### Impacts of CKD on PPV and outcomes

The current study reported a comparable percentage of patients who had VA improvement after PPV in both groups (46.4-54.3%), similar to previous work on PDR patients (49-75%)^28^ and PDR patients under dialysis (60.5%)^10^. In addition, we reported a similar high retinal attachment rate in both groups in the short-term follow-up (97.2%, 99.3%), similar to previously reported rates of 90.6-92.0%^28^; a 100% macular attachment rate in patients who underwent silicone oil removal, similar to previously reported rates of 66-88%^28^; a low occurrence of POVH in both groups (4,8), compared to previously reported rates of 15-32%^16,29,30^; and a zero occurrence of NVG in both groups, which has been reported to be 5.3-11.8%^31-33^ previously.

The poor VA outcome was known to be related to macular-involved TRD, incomplete PRP, severe neovascular elsewhere (NVE), the presence of NVI, tRD involving discs, poor pre-PPV VA, and the combination of tRD and a retinal tear^34^. The condition of high-risk PDR and the combination of retinal tears (32.9%, 39.7%) were similar in the two groups. However, patients in the impaired renal function group had a higher percentage of incomplete PRP, macular-involved tRD, and broader FVP that involved the disc (77.5%, 28.9%), which implied the risk of worse VA outcomes. However, both groups of patients achieved similar VA outcomes.

The goal of PPV in PDR patients is to clear the VH, release vitreoretinal traction, and complete PRP. There was no significant difference in the operation time, intraoperative laser points, or percentage of silicone oil tamponade between the two groups during PPV. However, we found that patients with impaired renal function had a higher percentage of subretinal fluid drainage, and severe intraoperative bleeding required endodiathermy.

Previous work showed that subretinal fluid could spontaneously resolve 2-12 months after PPV, and long-standing subretinal fluid is unrelated to visual prognosis^15-17^. Contrary to previous works focusing on the influence of subretinal fluid on visual recovery in PDR patients who underwent PPV, we focused on the presence of long-standing subretinal fluid, which may hold back the application of PRP and, in turn, increase the chance of postoperative NVI. Post-PPV NVG indicates poor visual prognosis in PDR patients^31-33,^ and complete intraoperative photocoagulation can effectively prevent post-PPV NVG^35^. Previous work performing PPV without subretinal fluid drainage on PDR patients showed that in a group of PDR patients with 41% of patients with macular-involved tRD and 48% of patients who had complete PRP before PPV, 7% of them developed NVG in the follow-up^36^.

Compared to previous work, our study had fewer patients with complete PRP before PPV (3/71) and a similar percentage of patients with macular-involved tRD (46.5%), but we had no NVG in the follow-up. If the subretinal fluid is left untreated, patients without complete PRP may develop NVG because it is difficult to perform additional PRP on the retina with extensive, long-standing subretinal fluid to correct retinal ischemia. Therefore, we performed subretinal fluid drainage in cases with a large area of tRD. In patients with impaired renal function, the higher percentage of patients with macular-involving tRD may explain the higher percentage of subretinal fluid drainage (45.1%, 22.1%). Our data suggested that subretinal fluid drainage in PPV in PDR patients, especially in patients with impaired renal function, should be performed considering the risk of NVG in addition to visual outcomes.

POVH is a common complication of PPV in PDR patients. Previous work indicated that pre-PPV IV anti-VEGF agents may lessen the chance of intraoperative bleeding and prevent POVH^37,38^. Although we had widely applied pre-PPV IV anti-VEGF treatment in both groups (81.7%,88.6%) and the FVP was more commonly observed as predominantly fibrotic (43.7%, 25.5%), the occurrence of severe intraoperative bleeding requiring endodiathermy was higher in the group of patients with impaired renal function (56.3%, 32.3%). It is known that extensive vitreoretinal adhesion is related to the aggression of NVE^39^. The high prevalence of extensive vitreoretinal adhesion in patients with impaired renal function (77.5%, 28.9%) may help to explain the high occurrence of severe intraoperative bleeding. The occurrence of POVH in both groups (4,8) was lower than previously reported as 15-32%^16,29,30^. Our results suggested that preoperative IV-anti-VEGF agents and the use of endodiathermy in cases with extensive FVP may be practical to lessen the chance of POVH.

### Limitations

Previous research showed that 44-72% of VH in PDR patients could resolve after IV-anti-VEGF agents in a follow-up of 16-52 weeks; only 7.04-33.0% of patients required PPV^40-43^. Since our study was a retrospective study with a relatively low percentage of patients who received IV-anti-VEGF agent after VH occurrence (6.5%, 7.5%), we could not show the influence of anti-VEFG agent injection on the resolution of VH and the chances of gaining a second PRP. The high prevalence of severe FVP in patients with impaired renal function in our patients suggested that the observation for VH absorption after IV-anti-VEGF agents in PDR patients combined with impaired renal function should not be too long in case of progression of tRD. The safe length of the observation period in patients with CKD after IV anti-VEGF agents should be further investigated.

We only had serum creatinine, blood urine nitrogen, and urine protein tests; we did not have a urinary albumin-to-creatinine ratio test. We could not show the influence of CKD with relatively normal renal function on the PDR characteristics or PPV details.

Incomplete PRP is related to the progression of PDR^44,45^ and the occurrence of VH^46^, POVH^27,^ and postoperative NVI ^31-33^. We reported a low prevalence of completion of PRP (3/71, 3/149) before PPV and a high number of laser points during the surgery (980, 1092). Nevertheless, due to the retrospective characteristics, we could not show the reason for incomplete PRP in our patients. The reasons for the incomplete PRP may be unawareness of DR or developing VH in high-risk PDR. Further investigation of the reason for incomplete PRP may provide more information on the influence of CKD on DR patients’ treatment before ophthalmic surgery.

## Conclusions

We showed that the prevalence of impaired renal function in PDR patients who underwent PPV was high, the awareness of the renal condition among PDR patients was low, and screening for CKD was required before PPV. We also found that PDR patients with impaired renal function had a high prevalence of extensive FVP, macular-involved tRD, and retinal vessel closure. In patients with extensive tRD, subretinal fluid drainage and photocoagulation are recommended to lessen the chance of NVG; moreover, pre-PPV anti-VEGF agents and endodiathermy in PPV could lessen the chance of POVH. In addition, patients with impaired renal function can achieve VA results and PPV outcomes comparable to those of patients with normal renal function after proper management during PPV.

## Data Availability

The dataset(s) supporting the conclusions of this article is(are) available in zhao, meng (2022), “PDR patients and vitrectomy”, Mendeley Data, V1, doi: 10.17632/gdkpjnjv45.1

https://doi.org/10.17632/gdkpjnjv45.1

